# Quantitative Estimation of Disruption in Social Contact Structure and its Effect in COVID-19 Spread in India

**DOI:** 10.1101/2020.04.27.20081620

**Authors:** Jay Naresh Dhanwant, V. Ramanathan

**Affiliations:** Department of Chemistry, IIT(BHU) Varanasi

## Abstract

In this paper we use the well-known SIR (Susceptible-Infected-recovered) epidemiology model for quantitatively estimating the impact of this disruption in the social contact structure of India and retrospectively estimate the number of COVID-19 cases in India by neglecting the single source event. Model predicts that around 32% of COVID-19 cases (as on April 14, 2020) are contributed by the single source event. Given this disruption of the social contact structure, the model shows that the country wide lockdown has been effective in bringing down the number of cases in India.

## Introduction

World is now grappling with the COVID-19 pandemic arising from the novel Corona virus. Several countries have adopted multi-pronged approach for containing the spread of the infection. With no specific vaccine or standard treatment protocols yet, the medical infrastructure of every country has forced them to adopt a sustainable strategy. As on date the virus has infected around 2.74 million people globally and this has resulted in the death of more than 191 thousand deaths in a total of 225 countries. Country-wide lockdown is one of the primary strategies that has affected the social contact structure and has checked spread of the infection albeit not at the desired levels. A one day country-wide lock down for 14 hours was observed in India pursuant to the call by the prime minister of India. Subsequently a three week lockdown in the whole of India was announced from March 24 until April 14, initially, which has now been further extended to May 03, 2020. Despite this call for social distancing pattern, there have been around 728 Indians have faced death and nearly 23,000 Indians have got infected [1]. During this lockdown the social structure was majorly disrupted by a single source event which has been attributed to the attendees of the congregation held at the Markaz Nizamuddin mosque in New Delhi. This congregation was held in the middle of March and subsequent country wide movement its attendees have resulted in disrupting the social distancing pattern of India as acknowledged in the press briefing by the Joint Secretary of the Ministry of Health and Family Welfare, Government of India. This event has been covered in the media by the name Tablighi Jamaat event.

In this paper we use the well-known SIR (Susceptible-Infected-recovered) epidemiology model for quantitatively estimating the impact of this disruption in the social contact structure of India. We have earlier used the customized SIR model and demonstrated the robustness of this model by validating with the pandemic data from Hubei and Italy [2]. We use the same customised model in to retrospectively estimate the number of COVID-19 cases in India by neglecting the single source event. In this way the impact of disruption in the social structure is estimated.

Epidemiological modelling work concerning the spread of COVID 19 has earlier been carried out using SIR and SIER models. One of the initial papers predicting the spread of COVID-19 in India was carried out SIR model by Singh and Adhikari [3]. This concerned the prediction of number of COVID-19 cases after the initial lockdown was lifted. They computed the basic reproductive ratio R0 and its time-dependent generalization based on case data, age distribution and social contact structure were also computed by them. They concluded then that the three-week lockdown (from March 24, 2020 – April 14, 2020) was insufficient to prevent resurgence. They suggested protocols of sustained lockdown with periodic relaxation besides predicting measures in reducing age-structured morbidity and mortality. One of the major drawbacks of their approach is that they have arbitrarily assumed the parameters in their model. Its another matter that their predictions for the number of COVID-19 cases for the middle of April is diametrically opposite to the reality.

Few other researchers have modelled COVID-19 spread using different machines learning algorithms. Some of the techniques that have been employed in this machine learning are ARIMA, polynomial fitting, neural networks for deep learning and exponential smoothing [4–9]. Neural Networks as well as third degree polynomial fit have been found to over-fit the data with a very high bias.

This is because the trend in the epidemic spread assumes different nature in different phases over time. A polynomial fit gives reliable results for short terms and deviates significantly with any change in social distancing pattern. Analytically solving the differential equations using a customised SIR model, described below is found to be more appropriate method.

In this work we have used a modified version SIR (Susceptible-Infected-Recovered) epidemic model. This model divides the entire population into three compartments, Susceptible, Infected and Recovered. Each compartment shares similar characteristics.

➢ S: Susceptible Population
➢ I: Infectious Population
➢ R: Recovered Population

The three categories are interrelated with each other with the parameters β and γ. The main drawback of this model is the presumption of the parameter β. In the modified approach that we have adopted, the parameters are not arbitrarily assumed rather we optimize these parameters through machine learning.

## Mathematical Model

Rate of change of Susceptible Population 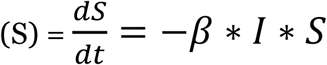

Rate of change of Infectious Population 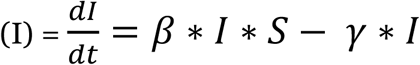

Rate of change of Recovered Population 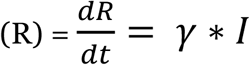

These equations were solved using Scipy and implemented in python [6]. The parameters β and γ are the key influencers and they are described below:

- β: extent of disease transmission of infection. β may be different for the same kind of virus in different societies. A lower value of β implies reduced social contacts in a society.
- γ: extent of recover in a specific time period
- α and 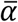: These are hidden parameters and not explicit in the above equations. They represent the measure of symptomatic cases + asymptomatic cases, respectively.

Given that there is no vaccine yet for the novel Corona virus yet and in the absence of standard medical protocols, the entire analysis revolves around social distancing patterns and hence the parameter β.

## Results and Discussion

It must be noted that the congregation, that impacted the social distancing pattern, had happened in the middle of March. Given that the incubation period of this virus, that is the number of days between contracting the virus and showing symptoms, is around 6 days. It was only after the 1^st^ April 2020 that news about this single source event started emerging and they were traced and tested which is caused a sudden spike in the number of infected cases in our country. This clearly shows that the attendees of the congregation were not tested in the first week of the lock-down 1.0 lest the number would have spiked early on, between March 23 and 30. In figure 1 we depict the ratio between the number of infected cases and the number of tests conducted. There are two trends shown in the figure. The blue trend line is the ratio taken for individual days and the orange trend line is ratio of the cumulative numbers. This ratio of new cases to total test, although too simplistic, yet indicates the need for more tests to be carried out if the ratio appears constant or increases over a period of time. In case this ratio trend line has even a light negative slope, it gives an early indication on the containment of the virus. From the figure we see that in both the trend lines, there is a significant spike in the first week of April, precisely on April 01 and 02, 2020. This is an early indication about the disruption in the social contact pattern.

**Figure 1:**
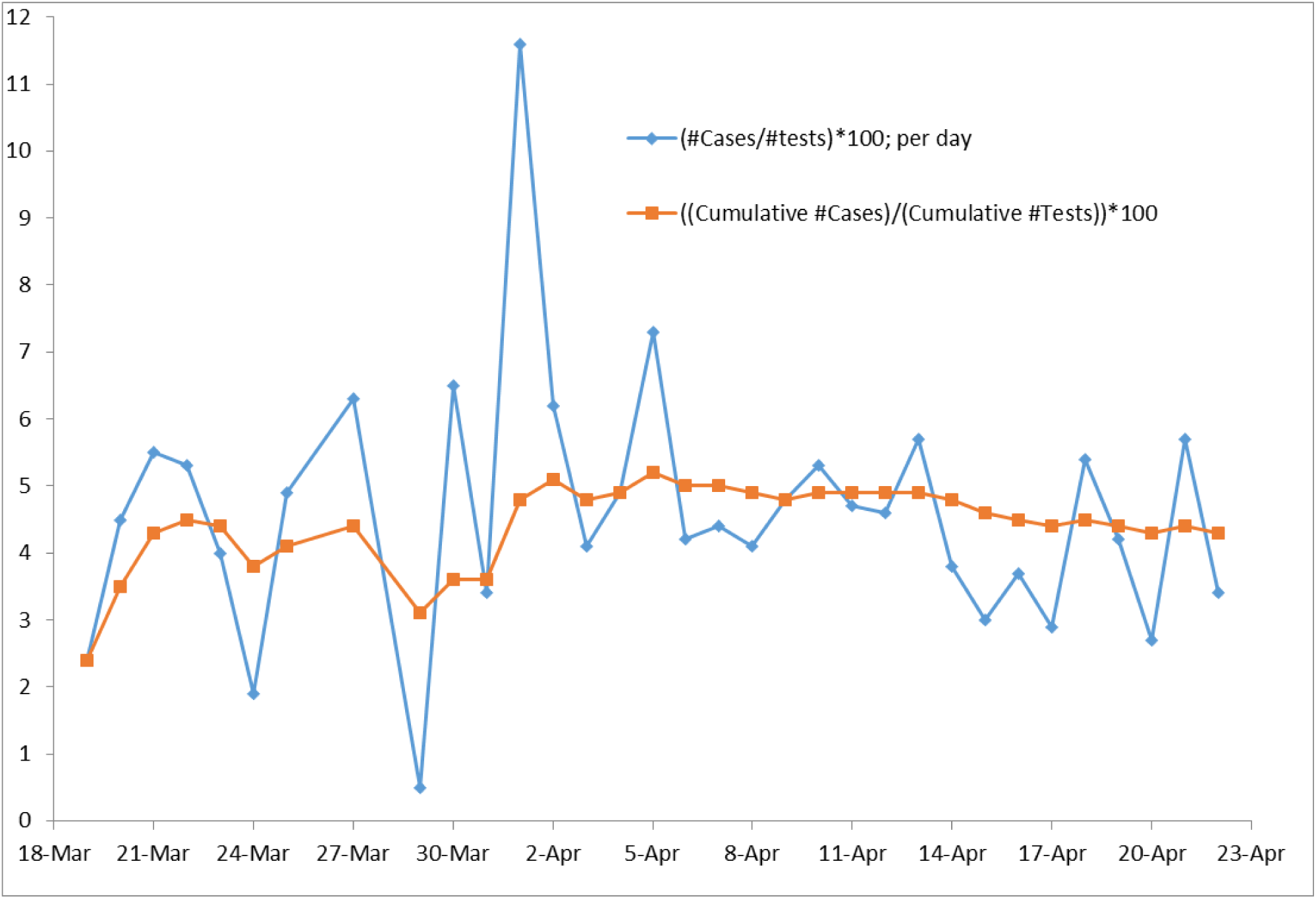
Ratio of number of positive cases to number of total number of tests, day-wise and cumulative.

We further aimed at quantifying the impact of the disruption in the social contact pattern for the period from 01/04/2020 till 14/04/2020 as this is the time span where the social disruption is revealed in the preliminary analysis. β value was learned from the data arising out of the above said period and In our earlier work [2] we showed that the value of β = 0.3536 was observed when the lockdown wasn’t imposed (before March 24, 2020), and β = 0.2627 was observed under the lockdown conditions (data from March 24 – March 30, 2020 used for machine learning). In Figure 2 the prediction from our model is depicted along with the actual in data as mall grey circles (real data taken from https://www.mohfw.gov.in/). The value of β is 0.3031 which is learned from the data accumulated in the first week of April. This figure once again highlights the robustness of our model where the predictions are in very good agreement with the real data.

**Figure 2:**
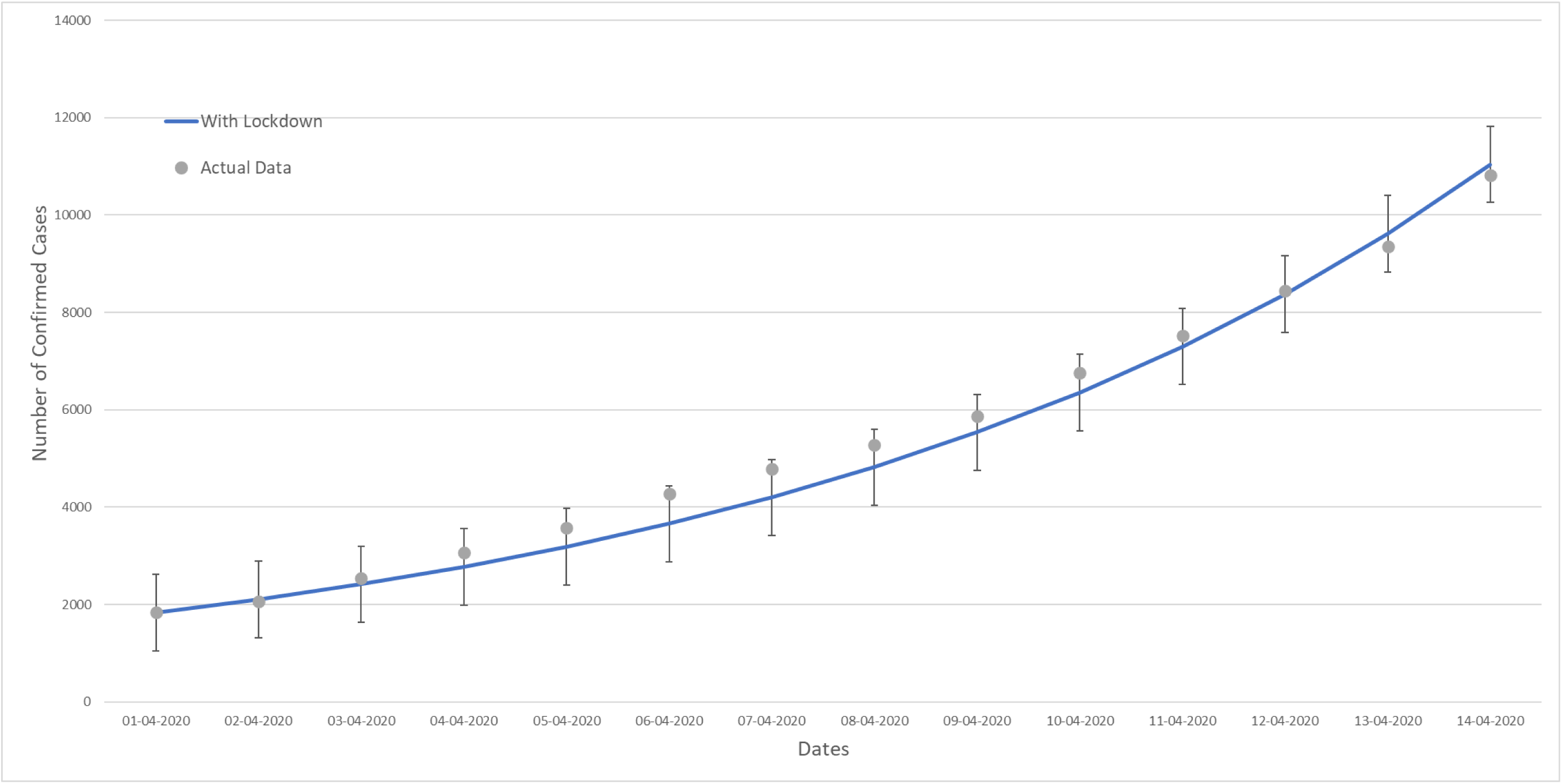
Proof of principle: validation of our model’s prediction with real data (Note: vertical lines indicate the standard deviation)

Further, the β = 0.3789 was used by spiking the β from the data before March 24, 2020 (β = 0.3526) and the trend was generated as a hypothetical case of no lockdown for the period of April 01, 2020 to April 14, 2020. β for the ‘no lockdown’ period was obtained by making a custom loss function which was minimized by using gradient descent on the loss function. The spike value was determined based on the increase in the average β value from March 24 – 30 2020 to the average β value in the first week of April. This is depicted as the orange trend line in Figure 3. Although the orange trend line is hypothetical, it still gives an approximated view of the impact of imposing a country wide lock down due to which the numbers of positive cases are orders of magnitude lesser in India.

**Figure 3:**
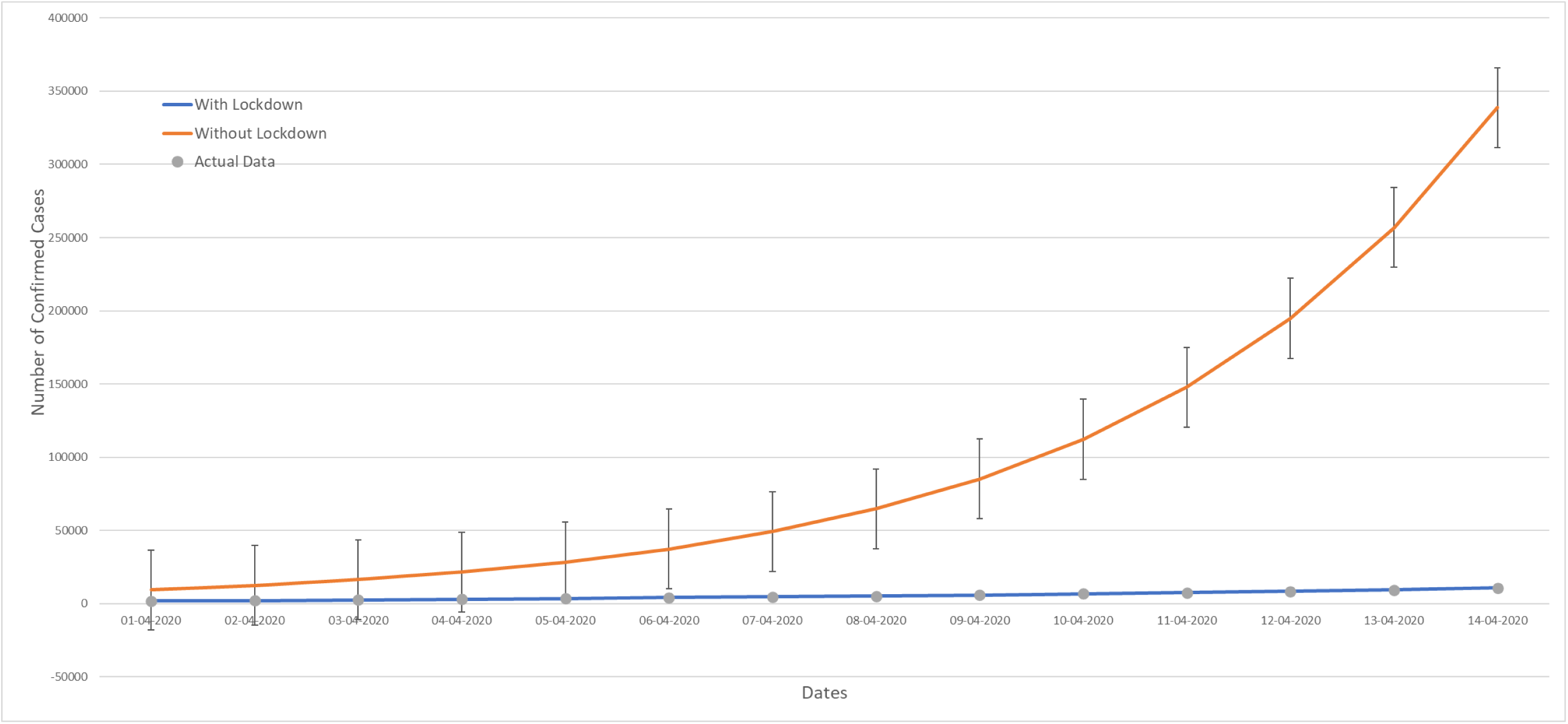
Impact of coutry wide lock down in bringing down the number of COVID-19 cases in India (Note: vertical lines indicate the standard deviation)

Having demonstrated that the predictions of our model are in close agreement with the real data, the impact of the disruption in social contact structure, the Markaz event, is presented in Figure 4. This is the prediction with the underlying assumption that the Markaz event had not happened. As mentioned above, the congregation happened in the middle March but the news about them started emerging only in the first week of April. Therefore the β = 0.2768 which is learned using the data from the first week of the lock down, March 24–30, 2020 is used for predicting the number of cases even for the period of lock down period from April 1 to 14. The blue trend line in the figure is the prediction by the model whereas the grey circles are the real data. From the figure it is indeed very apparent that the real data is significantly higher than the predictions. Actual number of infected cases as on April 14, 2020 is around 10,818 whereas our model predicts it to be around 7331 and this translates to that around 32.4% contribution of the Markaz event to the COVID-19 cases in India. This is in close agreement with the observation made by the officials in the Ministry of Health and Family Welfare, Government of India.

**Figure 4:**
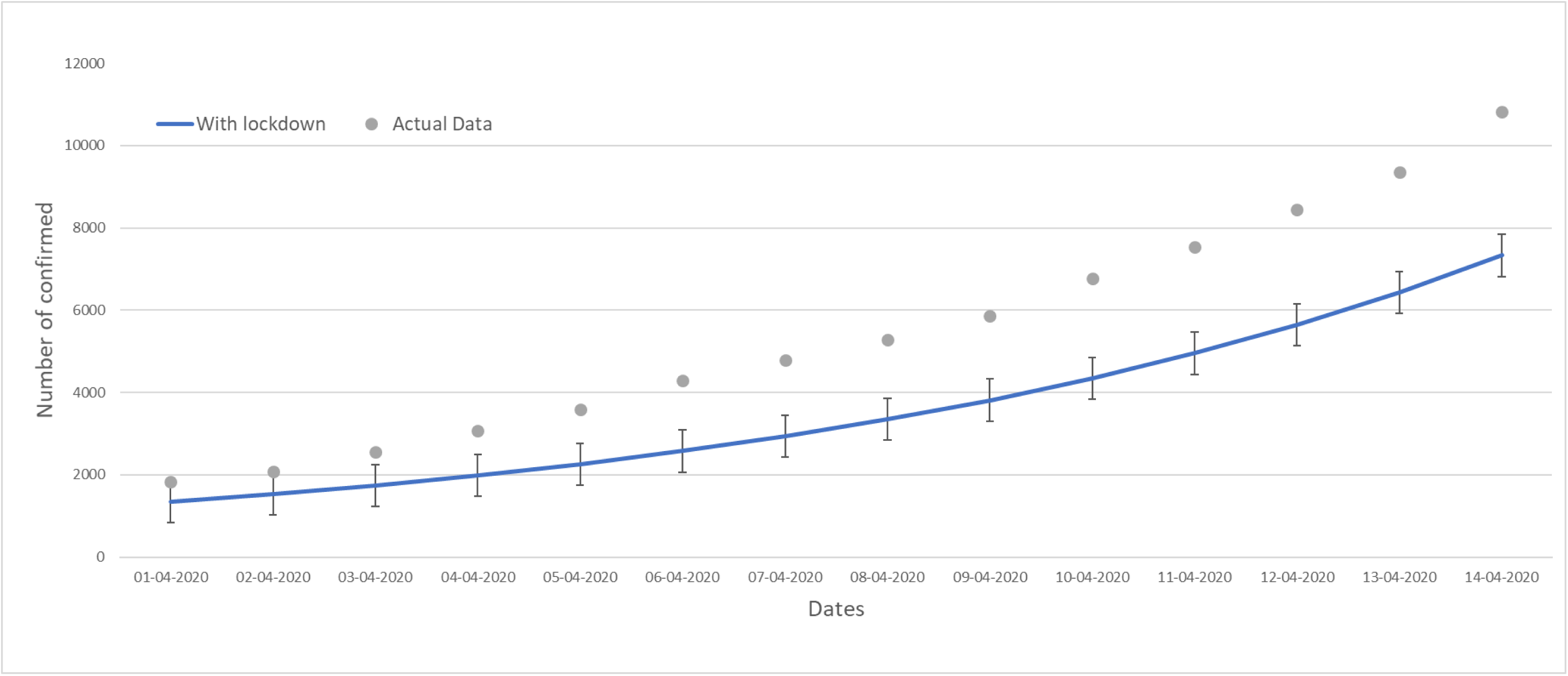
Impact of Markaz event in the number of COVID-19 cases in India. (Note: vertical lines indicate the standard deviation)

In Figure 5, the orange trend line depicts the hypothetical case of no lock down and no Markaz event by considering the beta value = 0.3526 which is learned from the data before March 24, 2020. The figure reveals that if there had not been any Markaz event and had there been no country wide lock down, the number of positive COVID-19 cases in India would have been nearly 0.1 million by April 14, 2020.

**Figure 5:**
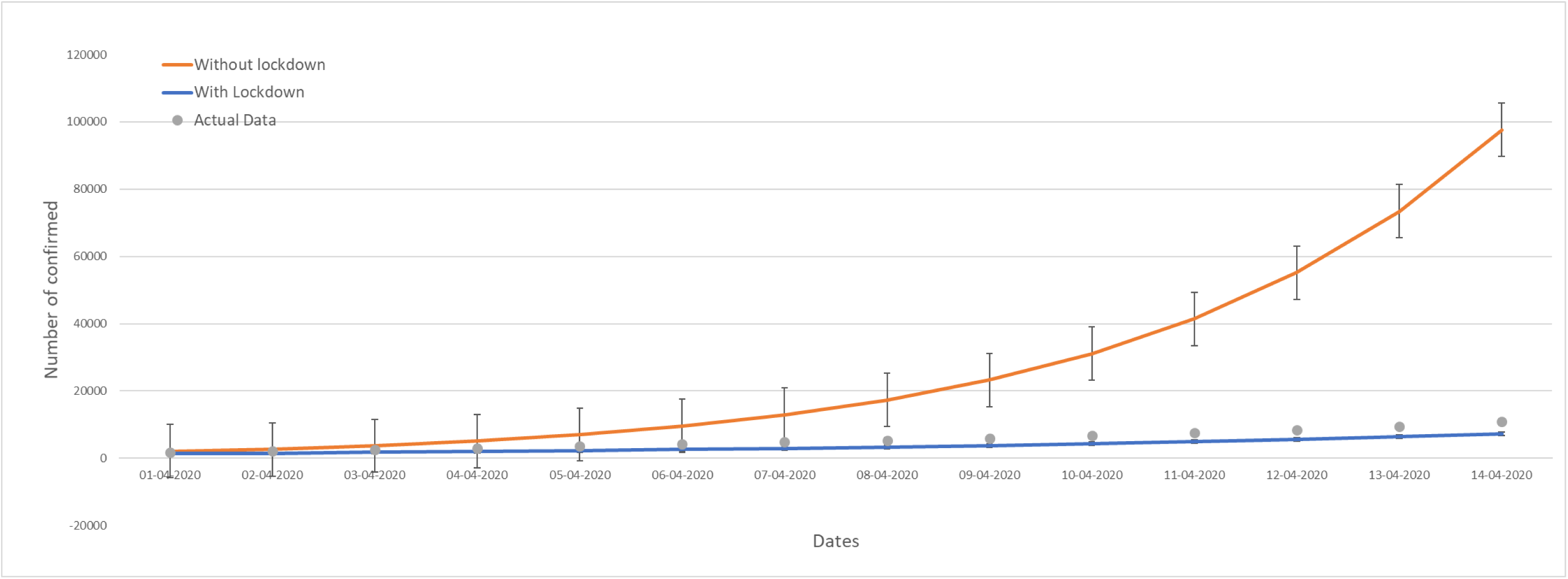
Predictions of our model in the absence of the lock-down and without considering the Markaz incident. (Note: vertical lines indicate the standard deviation)

Finally, the variation in the parameter beta for India is presented in Figure 6. These β values, as mentioned above, in the absence of vaccine and established medical protocol, measures the extent of the transmission of infection. Any change in this reflects the disruption in the social contact structure as well. β values presented in Figure 6 are learnt from a week long data. Lower value of beta directly corresponds to lower number of positively infected cases and vice versa. We see that the beta value spiked up to 0.3031 in the first week of April, owing to the Markaz event. Subsequently, owing to the restricted social contacting pattern through lock-down the value of beta then dropped to 0.2526. This trend once again reinforces the deleterious impact of the Markaz incident.

**Figure 6:**
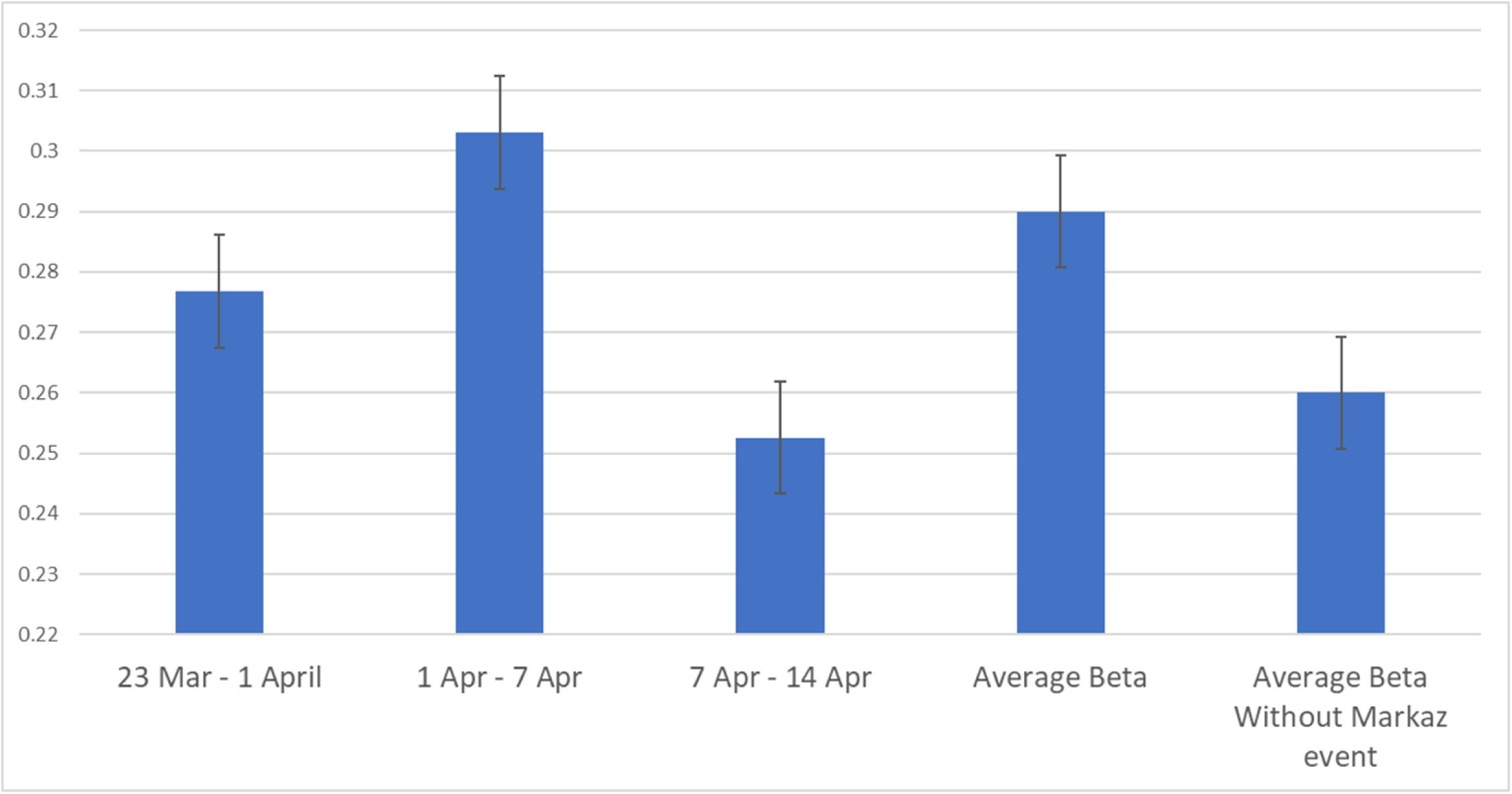
Social contact structures (with standard deviation) of India from March 23 till April 14.

## Conclusion

The customised SIR model with parameters learned using machine learning algorithms predict the spread of COVID-19 satisfactorily for India. Model predicts that around 32% of COVID-19 cases (as on April 14, 2020) are contributed by the Markaz event or what is also referred to as the single source event. Given this disruption of the social contact structure, the model shows that the country wide lockdown has been effective in bringing down the number of cases in India.

## Data Availability

Not applicable

## Sources

https://www.who.int/; https://www.mohfw.gov.in/

Dataset:

- Dataset was created and made live for open research by the Allen Institute for AI in partnership with the Chan Zuckerberg Initiative, Georgetown University’s Center for Security and Emerging Technology, Microsoft Research, and the National Library of Medicine - National Institutes of Health, in coordination with The White House Office of Science and Technology Policy.
- Another data set was made by Johns Hopkins University was used which was made live for an open research;

